# A multimodal investigation of altered cortical metabolism in patients with disorders of consciousness

**DOI:** 10.64898/2026.09.17.26363105

**Authors:** Niall W. Duncan, Likai Huang, Chia Lee, Yen-Chien Wu, Che-Ming Yang, David Yen-Ting Chen, Timothy J. Lane

## Abstract

Disorders of consciousness (DoC) are characterised by marked reductions in cerebral glucose metabolism, but the specific cellular and neurochemical processes driving this impairment remain unclear. To elucidate these mechanisms, a multimodal imaging approach combining [^18^F]-fluorodeoxyglucose positron emission tomography (FDG-PET) and magnetic resonance spectroscopy (MRS) was applied to the posterior medial cortex (PMC) in 15 DoC patients and 14 healthy controls. Regional FDG metabolic index (FDG-MI) values and MRS concentration estimates of glutamate plus glutamine (Glx), γ-aminobutyric acid (GABA+), total N-acetylaspartate (tNAA), total creatine (tCr), and total choline (tCho) were compared between groups and evaluated against Coma Recovery Scale–Revised (CRS-R) scores at baseline and follow-up (mean 127 days). Patients demonstrated reductions within the PMC in Glx, tCr, and tNAA concentrations, as well as a reduced Glx/GABA+ ratio. Local FDG-MI was substantially reduced in patients and correlated positively with tNAA and tCr. Furthermore, baseline tCho concentrations positively predicted long-term clinical recovery, as indexed by changes in CRS-R scores at follow-up. These findings suggest that macroscopic glucose hypometabolism in DoC may be linked to disrupted intracellular mitochondrial energy production pathways and a shift towards relative synaptic inhibition in the PMC. Moreover, the association between choline levels and behavioural recovery may point to the relevance of retained structural synthesis capacity for functional outcomes.

## Introduction

Disorders of consciousness (DoC), such as unresponsive wakefulness syndrome (UWS) and minimally conscious state (MCS), are conditions in which an individual displays long-term loss or severe limitation of conscious awareness but retains some degree of arousal fluctuations. These conditions present considerable clinical challenges in terms of treatment and prognostic determination (Edlow et al., 2021; Pavlov et al., 2024). At the same time, these clinical questions interface with wider questions around how consciousness arises from the human brain (Friedman et al., 2023; Gallucci et al., 2024). In particular, developing an understanding of the properties required for there to be the potential for conscious experience would be an important step for the development of treatments and prognostic markers (Schiff, 2024).

A change frequently reported in DoC patients is a global reduction in brain energy consumption, as indexed by fluorodeoxyglucose (FDG) PET. The magnitude of this reduction broadly follows consciousness levels measured with standard behavioural assessments, differing between patients diagnosed with UWS and those with MCS (Deruti et al., 2026; He et al., 2024; Liu et al., 2025; Stender et al., 2016). With the majority of glucose consumption in grey matter being dedicated to synaptic signalling, this graded decrease may indicate a progressive association between neural capacity and consciousness (Jamadar et al., 2025; Yu et al., 2018). At the same time, the metabolic state of the patient brain has been found in some studies to indicate the likelihood that there will be recovery to a higher level of consciousness in the future (Stender et al., 2016). Together, these results suggest that the capacity for a global conscious state is associated with ongoing neural signalling supported by a minimal metabolic state within the brain.

Neuroimaging measures of glucose metabolism present, however, only a partial view of cellular metabolism within the brain. They can indicate a change in overall metabolism but by themselves are limited in their ability to elucidate specific processes causing the alteration (Jamadar et al., 2025). In order to explore more deeply the metabolic changes occurring in DoC, we therefore employed a multi-modal imaging approach that combined FDG-PET with magnetic resonance spectroscopy (MRS). This imaging method allows the measurement of a limited set of specific chemicals within tissue, with the technique having been widely employed in studies of the human brain in health and disease (Duncan et al., 2014; Koush et al., 2022; McKiernan et al., 2023; Truong et al., 2021). Linking these two techniques may therefore allow the mapping of overall metabolic changes in DoC patients and relate that to more detailed cellular processes.

More specifically, MRS was used to estimate concentrations of glutamate and γ-aminobutyric acid (GABA) in the posterior medial cortex (PMC), along with levels of creatine, choline, and N-acetylaspartate (NAA). The PMC was selected as a target region for two reasons. Firstly, this region, as a component of the default mode network (DMN), has been consistently highlighted in studies of DoC (Crone et al., 2015; Qin et al., 2015b; Yang et al., 2024) and is often implicated in theoretical approaches to the neural correlates of consciousness (Albantakis et al., 2023; Ferrante et al., 2025). Secondly, this region generally allows for good MRS signal quality and so was hoped to provide more robust results from the DoC population. Glutamate and GABA were targeted as they respectively represent the major excitatory and inhibitory neurotransmitters in the brain. Measuring these in conjunction with glucose consumption would therefore better specify what transmission systems are altered in DoC, which cannot be directly established from FDG-PET measures alone. Creatine, choline, and NAA have multiple roles within the brain, including in cellular energy production, myelinogenesis, and neurogenesis. Identifying alterations in these and linking them to overall energy consumption may therefore highlight underlying disordered cellular processes in DoC.

## Methods

### Participants

Fifteen DoC patients and fourteen healthy controls (HC) were included in the analysis (see Table 1 for demographic details). A total of 63 patients underwent MRS scanning in this prospective observational study, but 48 of these were rejected due to poor data quality primarily due to head motion or the extent of tissue damage in the region of interest. Twelve of the included patients were diagnosed as UWS and three as MCS at the time of scanning. Clinical assessment was conducted by people trained in the use of the Coma Recovery Scale Revised (CRS) instrument (Giacino et al., 2004). Assessments were repeated at different times of day, with patients in a sitting posture where possible (DaCosta et al., 2025; Wang et al., 2020; Wilson et al., 2013). Follow-up assessments were also conducted (mean time to follow-up = 127 days, range = 68 - 195). One patient did not receive a follow-up assessment. Patient details are summarised in Supplementary table 1. None of the control participants had a history of neurological or psychiatric disorders. The study was approved by the TMU-Joint Institutional Review Board (N202204087 and N202109034). Written, informed consent was given by HCs and by family members or legal guardians of DoC patients. Other analyses from this sample are reported elsewhere (Han et al., 2024; Qin et al., 2018).

**Table 1:**
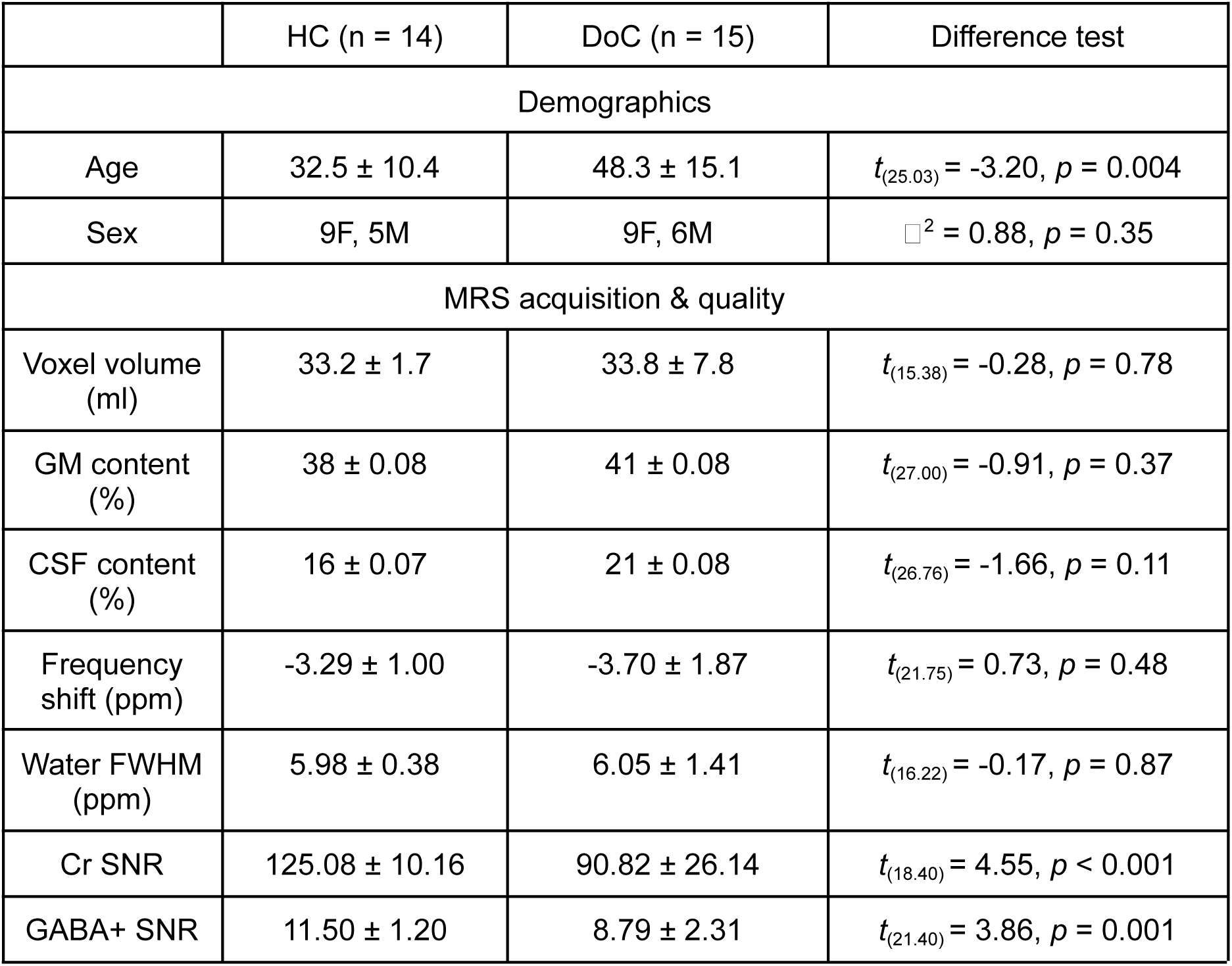
Participant characteristics, compared between groups. Cr = creatine; CSF = cerebrospinal fluid; DoC = disorders of consciousness patients; F = female; FWHM = full-width half-maximum; GABA+ = GABA+macromolecules; GM = grey matter; HC = healthy controls; M = male; SNR = signal-to-noise ratio.

|  | HC (n = 14) | DoC (n = 15) | Difference test |
| --- | --- | --- | --- |
| Demographics |  |  |  |
| Age | 32.5 ± 10.4 | 48.3 ± 15.1 | $t_{(25.03)} = -3.20, p = 0.004$ |
| Sex | 9F, 5M | 9F, 6M | $\chi^2 = 0.88, p = 0.35$ |
| MRS acquisition & quality |  |  |  |
| Voxel volume (ml) | 33.2 ± 1.7 | 33.8 ± 7.8 | $t_{(15.38)} = -0.28, p = 0.78$ |
| GM content (%) | 38 ± 0.08 | 41 ± 0.08 | $t_{(27.00)} = -0.91, p = 0.37$ |
| CSF content (%) | 16 ± 0.07 | 21 ± 0.08 | $t_{(26.76)} = -1.66, p = 0.11$ |
| Frequency shift (ppm) | -3.29 ± 1.00 | -3.70 ± 1.87 | $t_{(21.75)} = 0.73, p = 0.48$ |
| Water FWHM (ppm) | 5.98 ± 0.38 | 6.05 ± 1.41 | $t_{(16.22)} = -0.17, p = 0.87$ |
| Cr SNR | 125.08 ± 10.16 | 90.82 ± 26.14 | $t_{(18.40)} = 4.55, p < 0.001$ |
| GABA+ SNR | 11.50 ± 1.20 | 8.79 ± 2.31 | $t_{(21.40)} = 3.86, p = 0.001$ |

### MRI scanning

MRI data were acquired on a GE MR750 3-Telsa scanner using a standard 8-channel head coil. An FSPGR T1-weighted anatomical image was first acquired (TR = 8.2 ms, TE = 3.2 ms, flip angle = 8°, resolution = 1 x 1 x 1 mm3). This image was then used to locate the MRS voxel in the PMC. Voxels were targeted to align with the corpus callosum with the rear edge at the parietal-occipital boundary (see Figure 1). For patients, the location was made as close to this as possible while taking into account individual tissue loss and fluid-induced shifts within the skull. The target dimensions for the MRS voxels were 30 x 30 x 30 mm3, but this was adjusted depending upon participant anatomy and signal quality considerations. A MEGA-PRESS sequence was applied with the following parameters: TR = 1800 ms; TE = 68 ms; alternating ON/OFF editing with 14 msec editing pulses applied at 1.9 ppm (ON) and 7.46 ppm (OFF); 384 averages; 8 water unsuppressed acquisitions. MRI scans were conducted at the same time of day for HCs but varied for DoC patients due to clinical and logistical factors.

**Figure 1:**
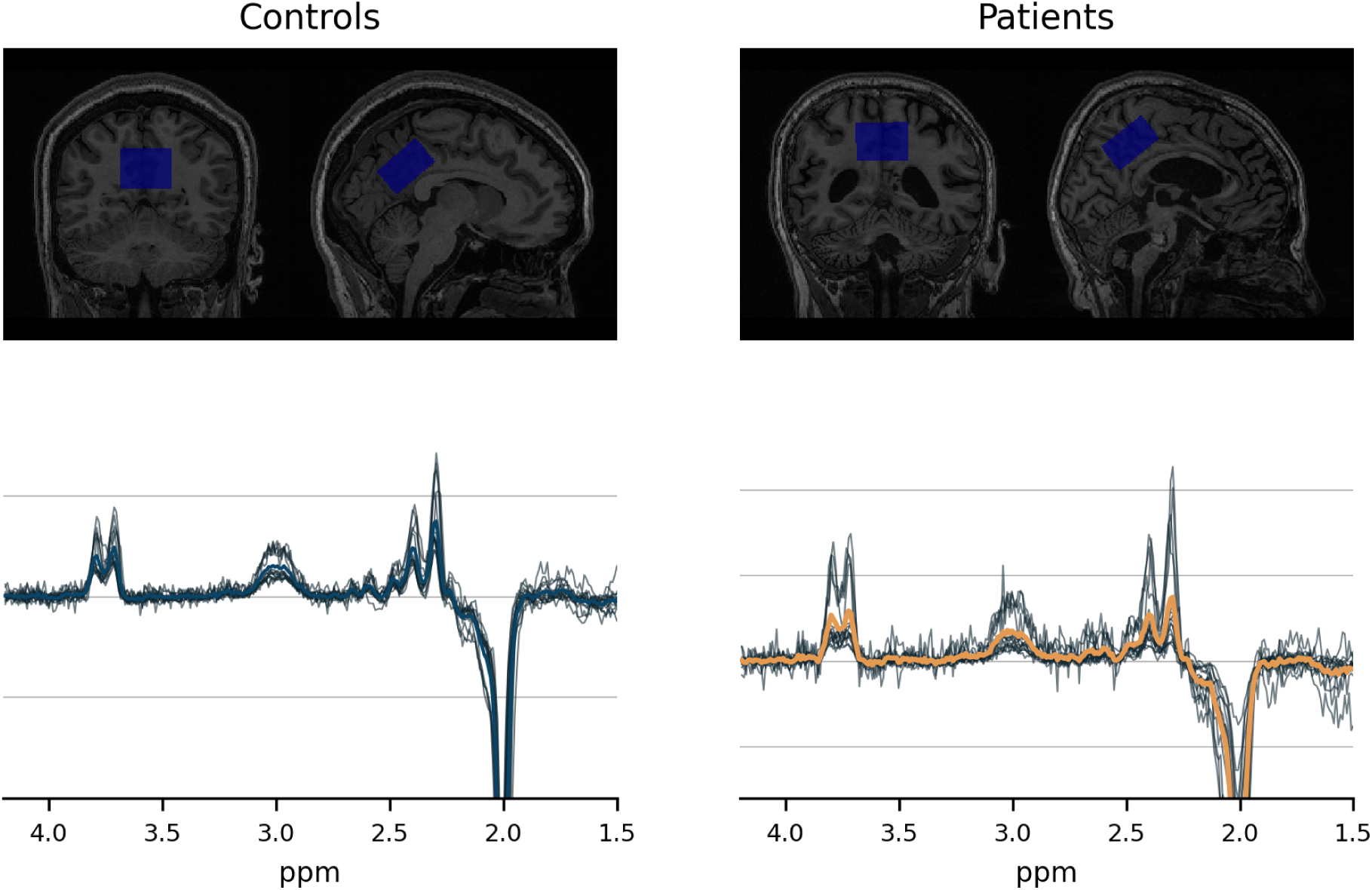
Example location of MRS voxel from a healthy control and patient (upper panels). MRS spectra from each group (lower panels). Individual spectra are shown along with the group average.

### MRI processing

MRS data were processed with the Osprey toolbox (version 2.7.1; Oeltzschner et al., 2020). GABA+macromolecule (GABA+) concentration estimates were made from the editing ON/OFF difference spectrum. Glutamate+glutamine (Glx), total choline (tCho), and total creatine (tCr) concentrations were estimated from the editing OFF spectrum. All metabolites were quantified in relation to the unsuppressed tissue water signal for the primary analysis. Quantification relative to tCr was also calculated for GABA+, Glx, tCho, and tNAA to test for the sensitivity of the results to reference type.

T1-weighted anatomical images were processed with FSL tools. They were segmented into grey matter, white matter, and cerebrospinal fluid (CSF) for use in the analysis of the data. They were also used to create a mask of non-brain tissue (head outline - brain) for FDG-PET analysis. Mask images were made for each participant’s MRS voxel in individual anatomical space. These were used to estimate the proportion of grey matter and CSF in the region.

### PET scanning and processing

FDG-PET data were acquired on a GE Discovery ST PET-CT scanner. HCs were asked to fast for eight hours prior to the scan session. [18F]-fluorodeoxyglucose (mean dose = 11.2 mCi ±1.4 SD) was administered intravenously, after which participants rested in a darkened room for 40 min. They were asked to lie with their eyes closed during this time. A 20-min scan was then conducted with the eyes closed. The same general protocol was used for DoC patients, including the delivery of the same instructions. DoC scans were scheduled for early morning to approximate an eight hour fast in a manner that did not interfere with their normal care.

Image reconstruction was done using an iterative process implemented in the manufacturer provided software. FDG uptake images were then aligned to the participant’s T1-weighted anatomical image. This was done by creating an individualised simulated FDG image by convolving grey matter density estimates from the anatomical image with an 8 mm FWHM Gaussian kernel (Baranova and Duncan, 2025) to which the FDG image was then aligned using ANTs tools. With the inverse of this alignment, the non-brain mask calculated previously was applied to the FDG-PET data and the trimmed mean (5th to 95th percentile) image value within this region used to normalise within-brain values to metabolic index (MI) values (Stender et al., 2016).

### Statistical analysis

Demographic and imaging properties were compared between groups with Welch’s t-test or chi-squared tests, as appropriate. Linear regression was used to test group differences in estimated MRS metabolites. Age, frequency shift, creatine SNR, water SNR, and grey matter and CSF volume were included in the model as potential confounding factors. The relationships between MRS metabolite estimates and CRS-R scores on the day of scanning and the change at follow-up were then tested in a similar fashion. FDG-PET MI values were compared between groups through linear regression, with age, grey matter, and CSF volumes included in the model to account for differences across participants. Finally, the relationships between MRS metabolite estimates and MRS region MI values were tested. FDR correction was used to control for multiple comparisons. Statistical analyses were conducted in Python and R.

## Results

### Participant characteristics

DoC patients were, on average, older than the HC group. The groups did not differ in gender composition. Patients had an average CRS-R score on the day of scanning of 5.6 (range = 1-12).

MRS voxel volumes did not differ between groups, nor did the estimated volume of GM and CSF within the voxels (see Table 1). Frequency shift across the acquisition was similar for both groups, as was the FWHM of the water unsuppressed scans water peak (Table 1). The estimated SNRs for both GABA and creatine were lower in DOC than in HC (Table 1). This difference in data quality concords with visual inspection of patient spectra (Figure 1).

### MRS - Group comparison

As shown in Figure 2 and Table 2, DoC patients showed reduced concentrations of Glx, tCr, and tNAA, compared to HCs. GABA concentrations did not differ between the groups but the estimated Glx/GABA+ ratio did. These group differences were also seen when using creatine as reference (the tCr group difference was not tested).

**Figure 2:**
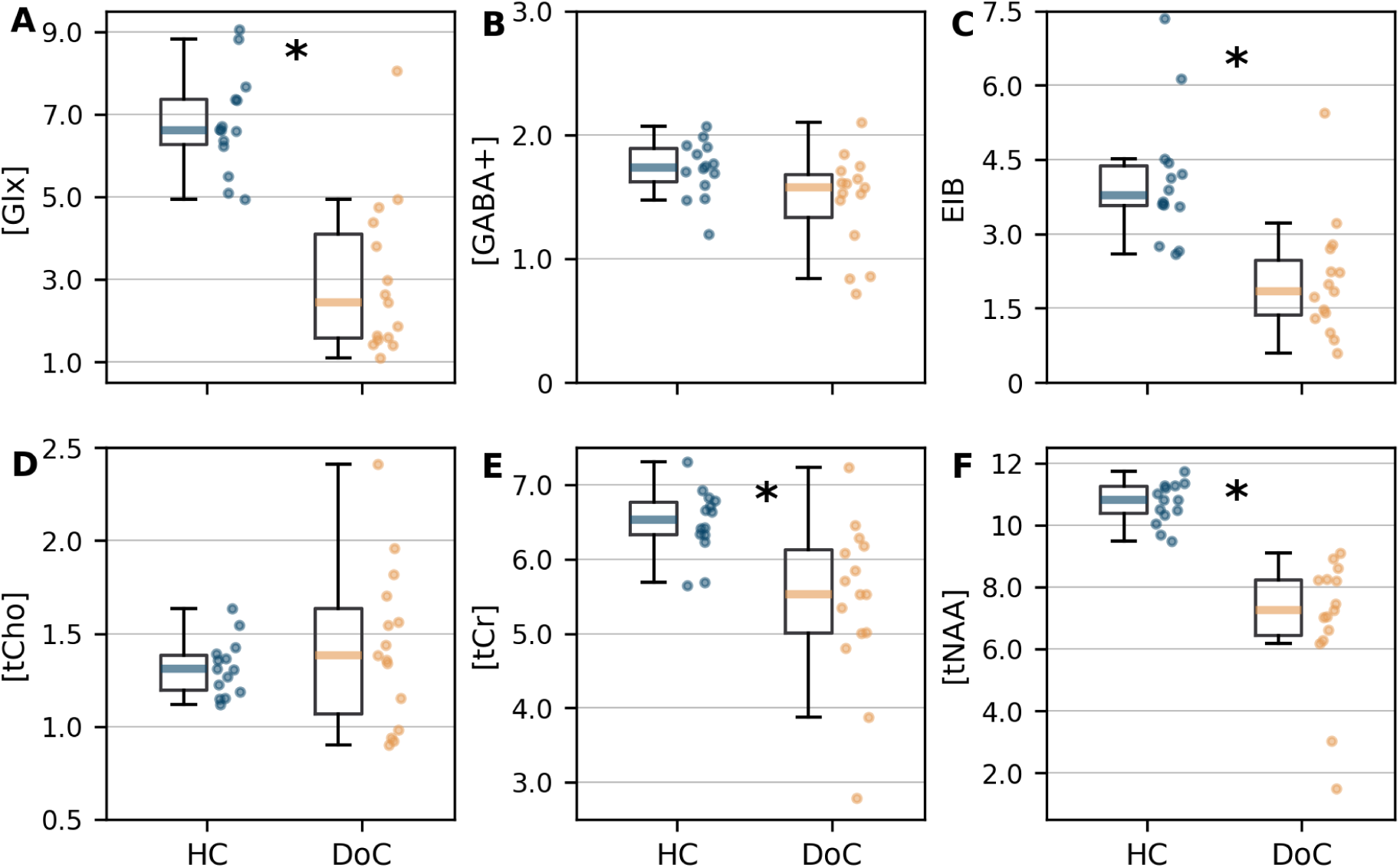
Comparisons of MRS estimates of brain metabolite concentrations between healthy controls (HC) and patients (DoC). * denotes p_FDR_ < 0.05. EIB = excitation/inhibition balance; GABA+ = GABA + macromolecules; Glx = glutamate + glutamine; tCho = total choline; tCr = total creatine; tNAA = total N-acetylaspartate

**Table 2:** Group differences in MRS metabolite estimates between healthy controls (HC) and patients (DoC). *ε^2^*effect sizes (and 95% confidence intervals) were estimated from the regression model. EIB = excitation/inhibition balance; GABA+ = GABA + macromolecules; Glx = glutamate + glutamine; tCho = total choline; tCr = total creatine; tNAA = total N-acetylaspartate; pFDR = adjusted p-value after FDR correction.

| | $\hat{\beta}$ | $t$ | $p$ | $p_{FDR}$ | $\epsilon^2$ [95% CI] |
| --- | --- | --- | --- | --- | --- |
| Glx | 2.61 | 3.91 | < 0.001 | 0.0024 | 0.39 [0.09 - 0.63] |
| GABA+ | 0.15 | 1.04 | 0.31 | 0.31 | 0.00 [0.00 - 0.17] |
| Glx/GABA+ | 1.40 | 2.36 | 0.028 | 0.042 | 0.17 [0.00 - 0.45] |
| tCho | -0.27 | -1.90 | 0.071 | 0.085 | 0.00 [0.00 - 0.38] |
| tCr | 0.81 | 2.65 | 0.015 | 0.029 | 0.22 [0.00 - 0.49] |
| tNAA | 3.08 | 5.77 | < 0.001 | < 0.001 | 0.60 [0.29 - 0.76] |

### MRS - Relationship with CRS scores

Uncorrected negative associations were found between CRS scores on the day of MRI scanning and both estimated Glx concentrations and the EIB. These did not survive correction for multiple comparisons (see Supplementary figure 1 and Supplementary table 2).

A positive association with tCho and the change in CRS scores between the day of MRI scanning and follow-up was observed (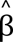 = 14.15, *t* = 3.86, *p* = 0.0084, *p_FDR_* = 0.05, *ε2* = 0.67 [0.06 - 0.86]; see Supplementary table 3 and Supplementary figure 2). This effect was also seen when using creatine as reference.

### FDG metabolic index

FDG metabolic index values within the MRS voxel were reduced in patients compared to controls ( 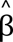 = 3.63, *t* = 6.21, *p* < 0.001, *ε^2^* = 0.62 [0.33 - 0.77]; Figure 3).

**Figure 3:**
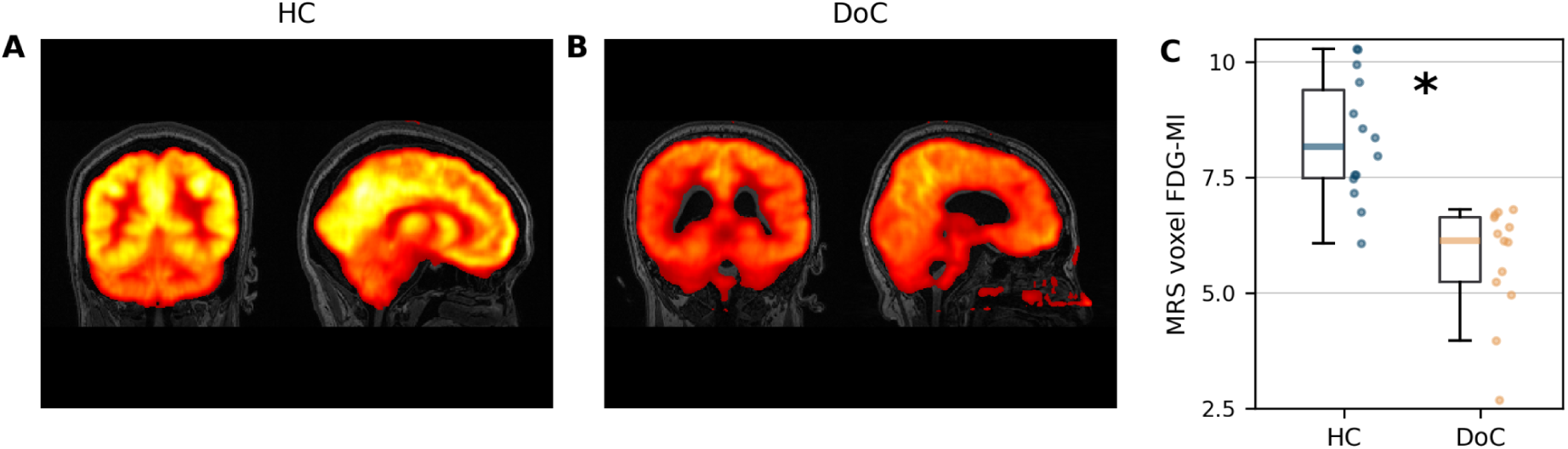
Example FDG-PET metabolic index (FDG-MI) images from (A) a healthy control and (B) a DoC patient. C) The metabolic index was reduced in DoC patients compared to controls. * denotes p < 0.05.

The within-voxel FDG-MI was associated with tCr and tNAA (Figure 4 and Table 3). An uncorrected association with Glx may also be noted. The tNAA association is also present when creatine is used as reference. This association is also robust to the removal of a potentially outlying patient (see Figure 4); however, the association with tCr is not (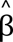 = 1.05, *t* = 1.71, *p* = 0.11, *ε^2^* = 0.09 [0.00 - 0.38]).

**Figure 4:**
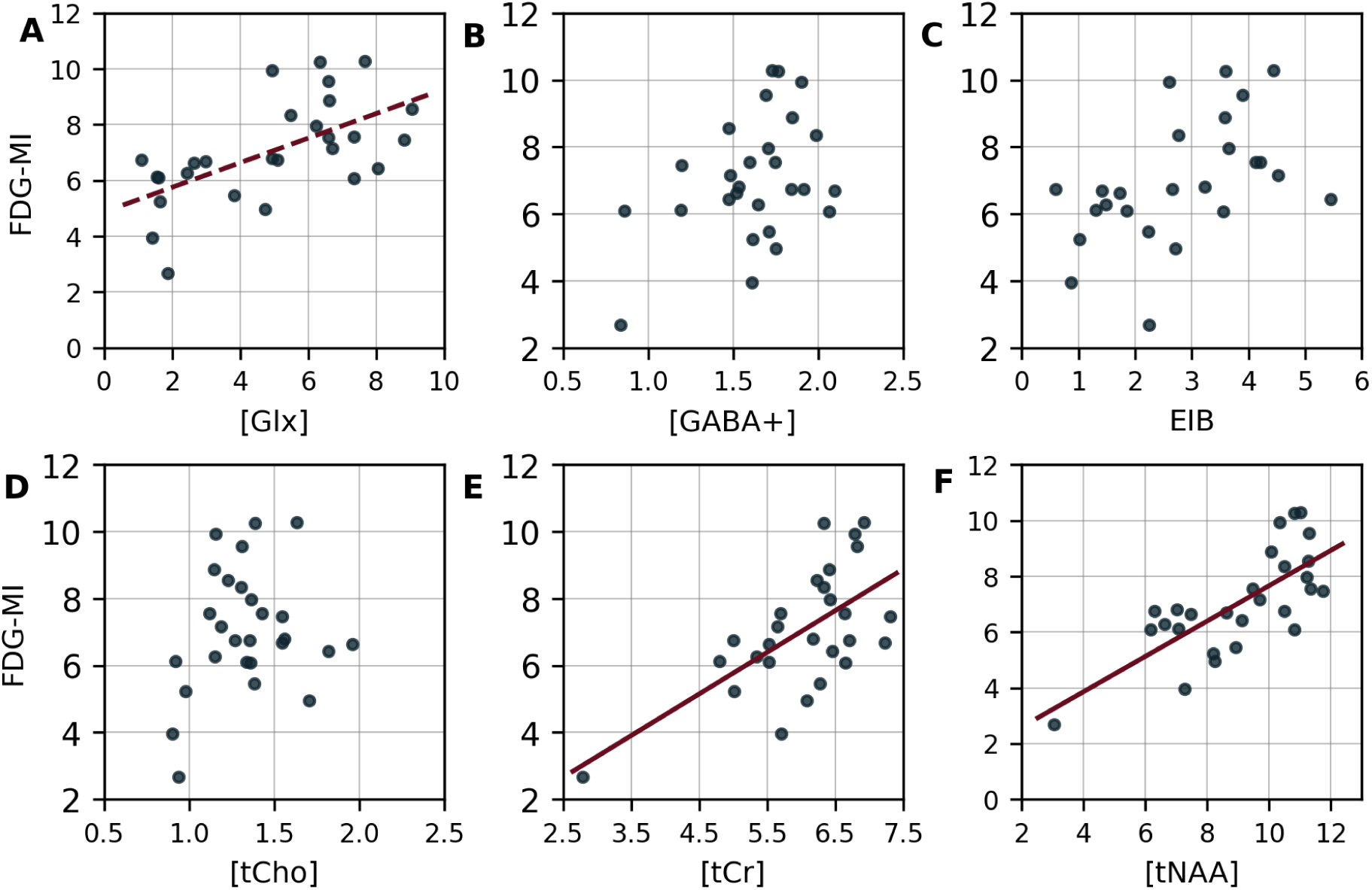
Associations between FDG metabolic index (FDG-MI) and metabolite estimates. Solid lines indicate an association that remains after FDR correction; dashed lines denote an effect that does not remain after FDR correction. The association of FDG-MI with tNAA remains when removing the potentially outlying patient but that with tCr does not (*p* = 0.11). EIB = excitation/inhibition balance; GABA+ = GABA + macromolecules; Glx = glutamate + glutamine; tCho = total choline; tCr = total creatine; tNAA = total N-acetylaspartate; FDG-MI = metabolic index calculated from FDG-PET data.

**Table 3:** Association between MRS metabolites and FDG metabolic index within the MRS voxel. *ε^2^* effect sizes (and 95% confidence intervals) were estimated from the regression model. EIB = excitation/inhibition balance; GABA+ = GABA + macromolecules; Glx = glutamate + glutamine; tCho = total choline; tCr = total creatine; tNAA = total N-acetylaspartate; *p_FDR_* = adjusted p-value after FDR correction.

| | $\hat{\beta}$ | $t$ | $p$ | $p_{FDR}$ | $\varepsilon^2$ [95% CI] |
| --- | --- | --- | --- | --- | --- |
| Glx | 0.51 | 2.13 | 0.046 | 0.09 | 0.15 [0.00 - 0.44] |
| GABA+ | 1.65 | 1.25 | 0.23 | 0.34 | 0.03 [0.00 - 0.27] |
| Glx/GABA+ | 0.32 | 1.02 | 0.32 | 0.39 | 0.00 [0.00 - 0.16] |
| tCho | 0.87 | 0.47 | 0.64 | 0.64 | 0.00 [0.00 - 0.00] |
| tCr | 1.33 | 2.65 | 0.016 | 0.047 | 0.23 [0.00 - 0.51] |
| tNAA | 0.94 | 4.35 | < 0.001 | 0.0021 | 0.47 [0.14 - 0.69] |

FDG metabolic index values within the MRS voxel were not associated with CRS scores on the day of scanning 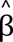 = -1.12, *t* = -1.66, *p* = 0.14, *ε^2^* = 0.16 [0.00 - 0.57]), nor were they associated with the change in CRS scores at follow-up (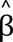 = 2.09, *t* = 2.05, *p* = 0.07, *ε^2^* = 0.26 [0.00 - 0.64]).

## Discussion

A multimodal imaging approach, combining MRS and FDG-PET, was used to investigate metabolic and biochemical alterations in the PMC of patients with DoC. Reductions in the concentration of tNAA and tCr were observed in patients compared to controls. Notably, whilst the absolute concentration of GABA+ was unchanged, the ratio of GABA+ to Glx was reduced. Furthermore, individual levels of tNAA and tCR were positively correlated with glucose consumption, as measured with FDG-PET, suggesting links between depressed energy metabolism at a macroscopic level and specific intracellular processes.

No difference in measured GABA+ levels was observed in DoC patients. At the same time, the ratio between Glx and GABA+ concentrations was shifted towards what could be interpreted as a relative inhibitory regime in DoC, with a reduction in excitatory signalling indicated by the lower Glx concentrations and inhibitory signalling remaining constant. This shift towards inhibition would fit with prior evidence linking inhibitory changes with a loss of consciousness (Dai et al., 2026) and with changes seen in anaesthesia (Zhang et al., 2009). One line of evidence for this connection is the effectiveness of GABAergic agents as general anaesthetics. Similarly, changes in inhibitory activity appear to be central to transitions between sleep and wake states. Functionally, it has been proposed that inhibitory balance in the brain is necessary for maintaining the level of integration and dissociation between networks that underlie the conscious state (Demertzi et al., 2022). This is supported by whole-brain modelling that integrates GABAergic tone to simulate functional activity patterns in both anaesthesia and DoC (Luppi et al., 2022).

The lack of a change in GABA+ observed here would appear to conflict with prior Flumazenil PET work that suggested overall reductions in GABA-A receptor availability in DoC (Qin et al., 2015a). However, although reductions across the brain as a whole were observed in that work, relative GABA-A receptor density was retained in PCC and precuneus in DoC. This suggests converging evidence within this region from the different methodologies and highlights potential variation in effects across the brain. More MRS-based studies of DoC that image multiple brain regions may be justified to investigate any such regional variation. The dorsal anterior cingulate cortex would be of particular interest as a change in GABA-A receptor density was highlighted there in the Flumazenil PET data (Qin et al., 2015a). A contrast between the posterior region studied here and anterior regions where GABA changes have been reported may be informative in the context of discussions of the relative importance of different brain areas for consciousness (Ferrante et al., 2025). It also concords with the need for the development of a multi-scale understanding of DoC, covering synaptic, circuit, and network levels, to support treatment and prognostic marker development (Schiff, 2024).

Although Glx levels can be interpreted as indicative of glutamatergic neurotransmission, glutamate (and glutamine, as its precursor) also plays a role in energy metabolism in the brain (Dienel, 2019). Separating out these functions is challenging, not least because a large proportion of energy within neurons is dedicated to transmitter-based signalling (Yu et al., 2018). Here we see tentative evidence for a correlation between measured Glx concentrations and FDG uptake, suggesting that a connection between the measured Glx and energy metabolism is present. The available data does not, however, allow us to discriminate between an association due to glutamate’s role in energy production or energy use for its synaptic release.

Clearer evidence for altered intracellular energetic processes in DoC comes from the clear reduction of the FDG metabolic index, coupled with correlated reductions in tNAA and tCr. Although commonly connected with lipid production (Chakraborty et al., 2001; Moffett et al., 2013), the synthesis of NAA has also been suggested to act as a buffer for aspartate arising from α-ketoglutarate production within mitochondria (Puthillathu et al., 2026). Removal of this product may be necessary for continuous synthesis of α-ketoglutarate through the GOT2 pathway for input into the TCA cycle. Notably, glutamate is the substrate for GOT2 synthesis of α-ketoglutarate meaning there is a plausible connection between changes in both Glx and tNAA in DoC. This is supported by a strong correlation between these substances in the current patient group (Spearman’s ⍴ = 0.79, *p* < 0.001). Creatine (plus phosphocreatine) also plays a role in energy production and distribution processes within the cell (Lowe et al., 2013). In particular, it provides ADP to the mitochondrial ATP synthesis pathway and is the precursor for ATP creation within the cytosol when required (Wallimann et al., 1992). Taken together, the observed reductions in these metabolites may provide converging evidence of altered mitochondrial energy production in DoC (Tao and Fujisawa, 2026), potentially contributing to an inhibited energy state that is incapable of supporting conscious awareness (Chen and Zhang, 2021; Stender et al., 2016).

The highlighted metabolites (tNAA and tCr) also play important roles in the maintenance of cellular integrity and in regeneration, both through contributions to relevant pathways and through their role in energy production (Chakraborty et al., 2001; Correale, 2025; Gallo, 2020; Tokarska-Schlattner et al., 2012). Also important in the control of cell structure is choline, which is necessary for the formation of lipid membranes and myelin (Skripuletz et al., 2015; Zeisel et al., 1991). Although we do not see an association between tNAA, or tCr and patient’s consciousness level at the time of scanning, we do see a positive association between tCho and the change in consciousness scores at follow-up. This may point to there being a required level of retained structural synthesis potential within the brain to support improvement in consciousness over time. Were this to be the case, then targeting such processes may lead to effective interventions or may act as prognostic markers.

### Limitations

The primary limitation of the work is the relatively low number of patients who had usable imaging data. This affects the statistical power available and may influence the robustness of the findings. Replication in an independent sample is required. The exclusion of a large proportion of patients also means that there may be some bias in the type of patients ultimately included in the analysis (i.e., a subset who did not show large movements in the scanner and where damage to the region of interest was not too extensive). Patient sedation was not used in this study but this may be a useful approach in future ones in order to minimise in-scanner motion. A second limitation is that MRS voxel localisation in patients with extensive brain tissue damage is challenging. This can reduce the anatomical specificity of the results. Higher field MRI allows smaller voxels to be used for MRS acquisition and so the use of such scanners would be advisable for future investigations. This would also allow for higher measurement accuracy and metabolite discrimination.

### Conclusion

In line with prior studies, we find that energy metabolism within the PMC is reduced in DoC patients. This reduction was then shown to be connected to changes in specific intracellular metabolites. These metabolic changes suggest an alteration to the energy production pathways within the PMC. Such changes may mean that a minimal level of neuronal activity necessary for conscious awareness cannot be supported in these patients. At the same time, an association between improvements in consciousness level over time and metabolites involved in cellular integrity and regeneration point to a need for adaptive intracellular processes to be retained to support recovery. These findings remain preliminary at this point but highlight potential future directions for research and treatment development.

## Acknowledgments

The authors would like to thank all the support staff at Shuang-Ho Hospital for their care of patients and assistance with the research. They would also like to express their gratitude to the families of the patients that were involved in the study. This work was supported by grants to TJL from the Taiwan National Science and Technology Council (105-2632-H-038-001-MY3, 106-2410-H-038-003-MY3, 111-2410-H-038-012, 114-2811-B006-072, and 115-2811-B-006-048), as well as by charitable donations administered through the Taiwan Ministry of Health and Welfare (Registration number: T063), designated for the Brain and Consciousness Research Centre, Shuang Ho Hospital. NWD also acknowledges support from the Taiwan National Science and Technology Council (113-2423-H-038-002-MY3; 110-2628-H-038-001-MY4).

## Data availability

Data necessary for reproducing the results and figures presented here are available at https://doi.org/10.5281/zenodo.22782988. Raw imaging data is not available due to ethical limitations on the sharing of patient data.

## Author contributions

NWD: Conceptualisation; Formal Analysis; Methodology; Visualisation; Writing - original draft

LH: Investigation; Project administration

CL: Data Curation; Investigation; Project administration

CMY: Investigation; Resources;

YCW: Investigation DYC: Investigation

PQ: Conceptualisation; Investigation; Methodology

TJL: Conceptualisation; Funding acquisition; Investigation; Writing - original draft All authors reviewed and approved the final manuscript.

## Supplementary materials

**Supplementary Figure 1:**
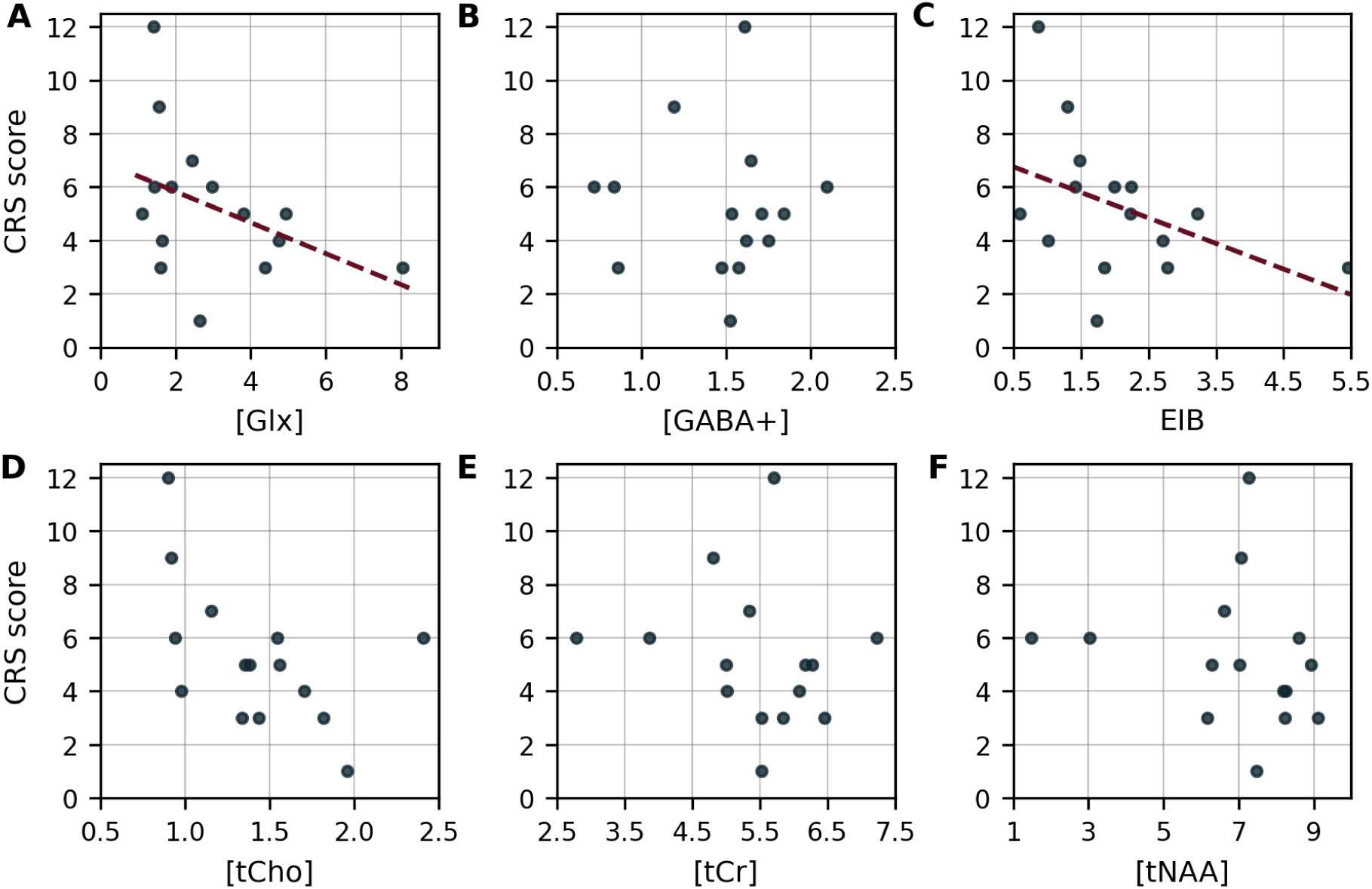
Associations between MRS metabolite measures and Coma Recovery Scale (CRS) scores at the time of scanning. Dashed lines denote an effect that does not remain after FDR correction.

**Supplementary Figure 2:**
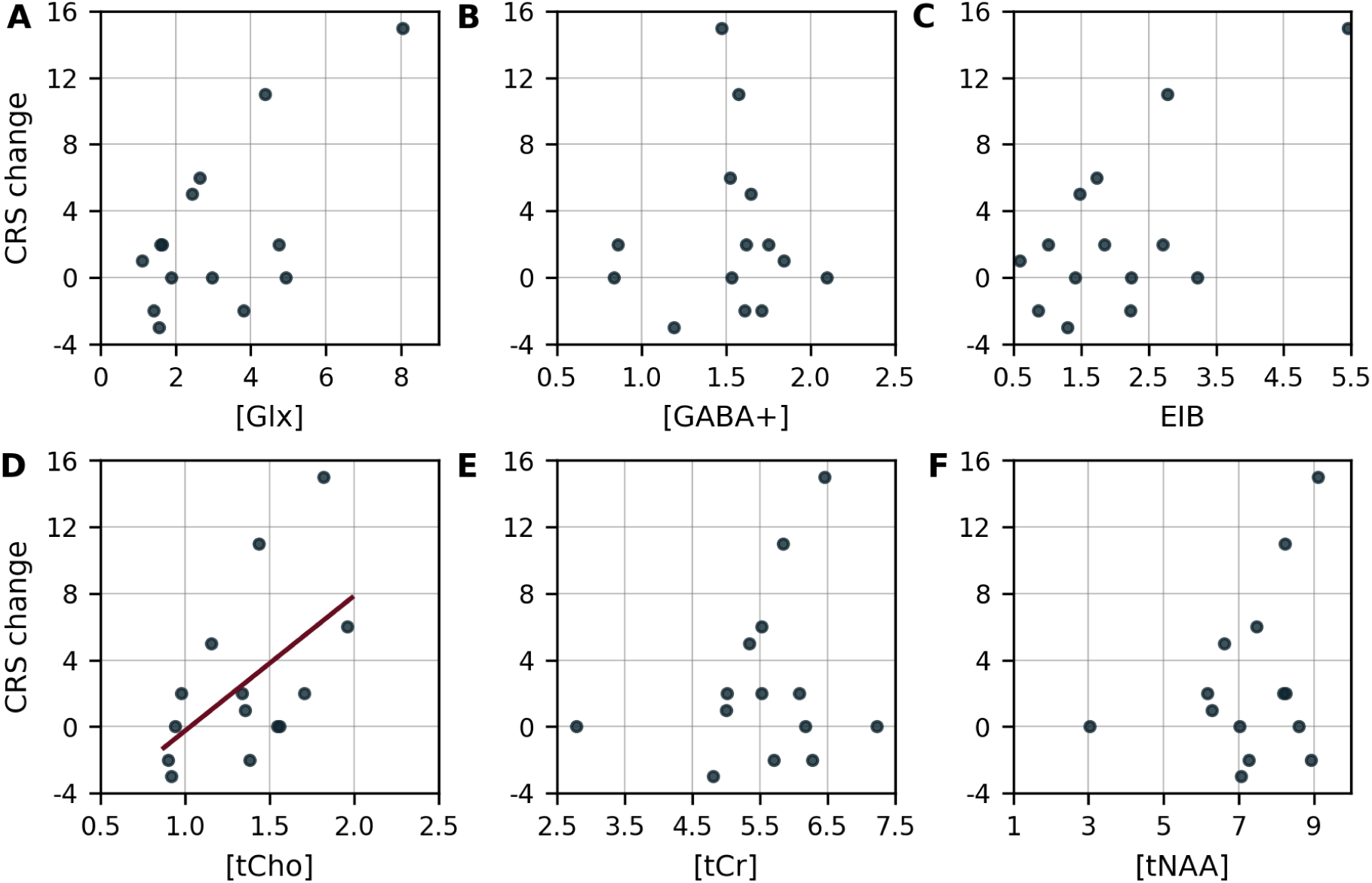
Associations between MRS metabolite measures and the difference in Coma Recovery Scale (CRS) scores between the time of scanning and at follow-up. Solid lines indicate and association that remains after FDR correction.

**Supplementary Table 1:** Patient details. The decade within which patient ages fall are given to reduce individual identifiability. CVA = cerebro-vascular accident; TBI = traumatic brain injury; UWS = unresponsive wakefulness syndrome; MCS = minimally conscious state; CRS = coma recovery scale.

| Patient number | Age | Sex | Aetiology | CRS at MRI | Diagnosis | CRS at follow-up | Time to follow-up (days) |
| --- | --- | --- | --- | --- | --- | --- | --- |
| 1 | 30s | M | CVA | 5 | UWS | 3 | 98 |
| 2 | 70s | M | CVA | 9 | MCS | 6 | 145 |
| 3 | 20s | M | TBI | 4 | UWS | 6 | 138 |
| 4 | 50s | M | TBI | 4 | UWS | 6 | 166 |
| 5 | 60s | F | TBI | 3 | UWS | 5 | 122 |
| 6 | 50s | F | CVA | 1 | UWS | 7 | 115 |
| 7 | 50s | F | TBI | 5 | UWS | 6 | 108 |
| 8 | 50s | M | TBI | 12 | MCS | 10 | 128 |
| 9 | 50s | M | TBI | 5 | UWS | 5 | 142 |
| 10 | 30s | F | unknown | 6 | UWS | - | - |
| 11 | 40s | F | TBI | 6 | UWS | 6 | 195 |
| 12 | 50s | M | CVA | 6 | UWS | 6 | 68 |
| 13 | 20s | M | TBI | 3 | UWS | 18 | 101 |
| 14 | 50s | F | CVA | 12 | MCS | 5 | 107 |
| 15 | 20s | M | TBI | 3 | UWS | 14 | 154 |

**Supplementary Table 2:**
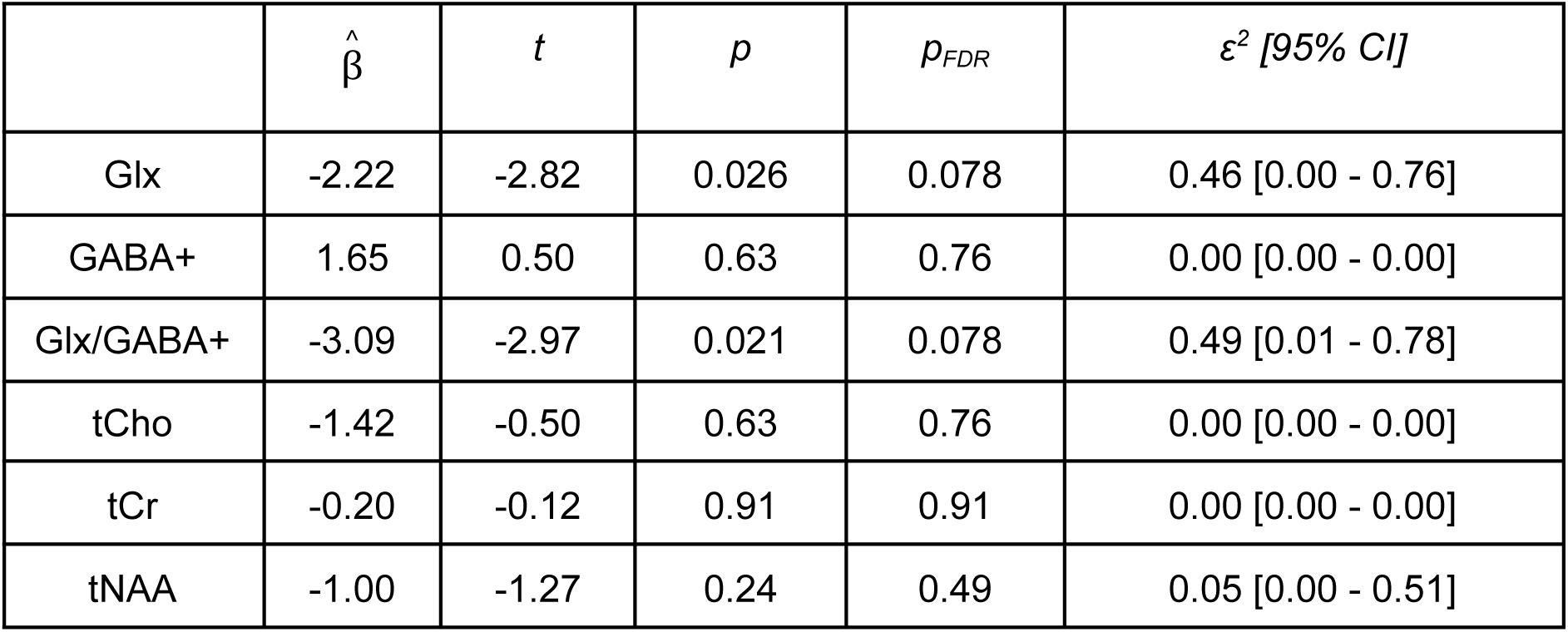
Association between MRS metabolites and CRS scores on the day of MRI scanning.

| | $\hat{\beta}$ | $t$ | $p$ | $p_{FDR}$ | $\epsilon^2$ [95% CI] |
| --- | --- | --- | --- | --- | --- |
| Glx | -2.22 | -2.82 | 0.026 | 0.078 | 0.46 [0.00 - 0.76] |
| GABA+ | 1.65 | 0.50 | 0.63 | 0.76 | 0.00 [0.00 - 0.00] |
| Glx/GABA+ | -3.09 | -2.97 | 0.021 | 0.078 | 0.49 [0.01 - 0.78] |
| tCho | -1.42 | -0.50 | 0.63 | 0.76 | 0.00 [0.00 - 0.00] |
| tCr | -0.20 | -0.12 | 0.91 | 0.91 | 0.00 [0.00 - 0.00] |
| tNAA | -1.00 | -1.27 | 0.24 | 0.49 | 0.05 [0.00 - 0.51] |

**Supplementary Table 3:**
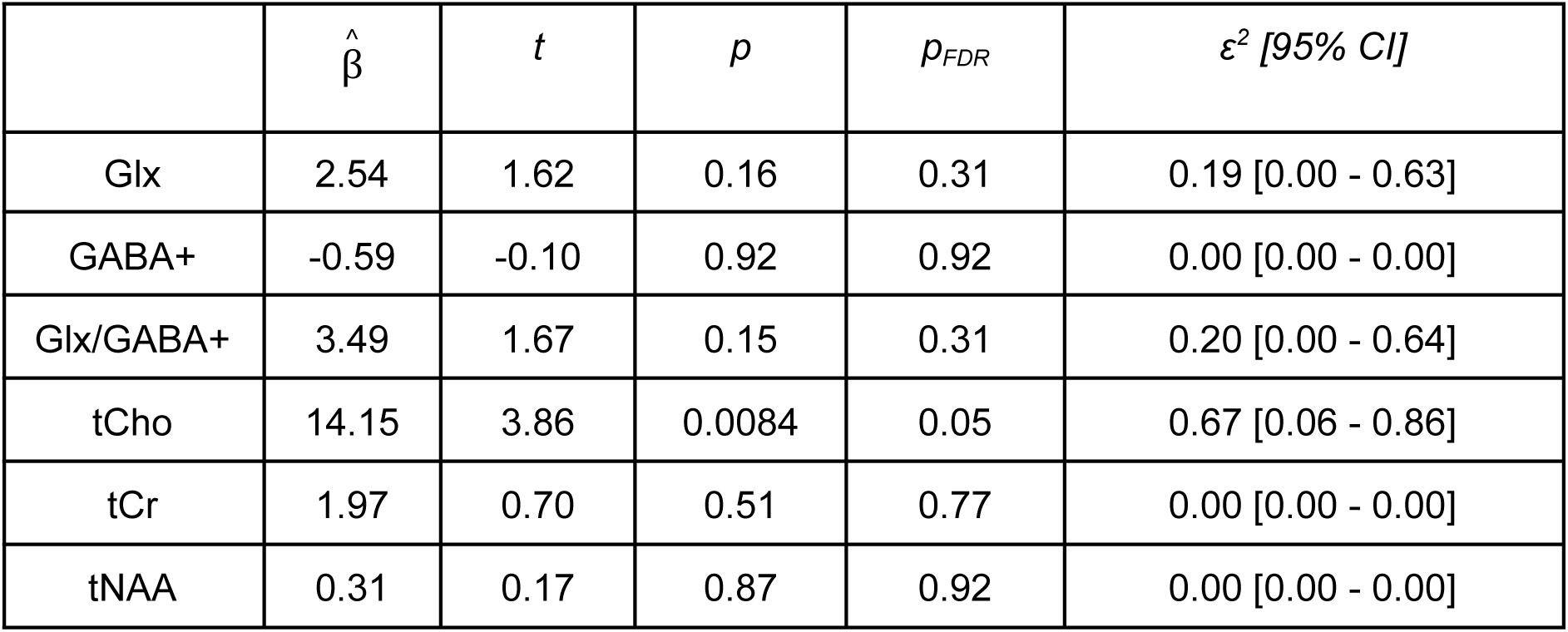
Association between MRS metabolites and changes in CRS scores between the day of MRI scanning and follow-up.

